# A Stage-Ordered Multi-Omic Continuum Underlies Cardiovascular-Kidney-Metabolic Syndrome and the Protective Association of Cardiovascular Health

**DOI:** 10.64898/2026.08.12.26360091

**Authors:** Yiwei Zhang, Xianglian Cai, Yanjun Zhang, Xiaoqin Gan, Yu Huang, Dan Chen, Xiaolong Liang, Yu Wang, Yuanyuan Zhang, Xianhui Qin

## Abstract

**Background and aims:** Cardiovascular-kidney-metabolic (CKM) syndrome stages confer graded CVD risk, but the underlying stage-specific molecular mechanisms remain undefined.

**Methods:** In 355,724 UK Biobank participants (median follow-up 13.5 years), we mapped CKM stages (0-3) to incident CVD. Using proteomics (n=37,785) and metabolomics (n=190,112), we identified stage-specific biomarkers via LASSO and XGBoost-SHAP. Mediation analyses were performed to quantify the proportion of the CKM–CVD association that was statistically accounted for by these biomarkers. The proportion of the protective association between cardiovascular health (Life’s Crucial 9 [LC9]) and incident CVD that was mediated by the same molecules was quantified.

**Results:** CVD risk increased across CKM stages. Beyond 11 pan-stage proteins (e.g., RTN4R,LEP) and 29 pan-stage metabolites (e.g.,GlycA), stage-specific molecular signatures emerged, whose pathway enrichment revealed a shift from metabolic/extracellular matrix dysregulation (Stage 1) to inflammation (Stage 2) to hypoxia/fibrosis (Stage 3). The proportion of the CKM–CVD risk association statistically accounted for by these molecules shifted accordingly: ADM (42.9%) in Stage 1, FABP4 (24.6%) in Stage 2, and HAVCR1 (28.0%) in Stage 3. High CVH (LC9≥80) was associated with approximately 80% lower CVD risk in Stages 0–2; a proportion of this protective association was statistically accounted for by the same stage-specific molecules.

**Conclusions:** These findings reveal a stage-ordered molecular continuum—from ECM remodeling to inflammation to fibrosis—that redefines CKM-driven CVD risk, and the strong protection of high CVH in early stages was statistically accounted for in part by these stage-specific molecules, generating the hypothesis that CVH may reduce risk through these modifiable pathways and providing a molecular framework for future stage-adapted intervention trials.

## Introduction

Cardiovascular disease (CVD) risk is substantially amplified by the interplay of metabolic, renal, and diabetic pathologies [1–3]. The American Heart Association introduced the cardiovascular-kidney-metabolic (CKM) syndrome framework, defining progressive stages (0–3) that capture the cumulative burden of these interacting conditions [4]. The 2026 multisociety clinical practice guideline subsequently standardized the implementation of this staging system in routine care [5]. Most recently, the 2026 Chinese Expert Consensus on CKM Syndrome, jointly developed by 121 experts across cardiology, nephrology, endocrinology, and related disciplines, further adapted this framework to Chinese populations and, critically, prioritized the discovery of high-sensitivity, stage-specific biomarkers to advance mechanistic understanding [6]. However, the molecular architecture that translates CKM burden into excess CVD risk remains undefined [4,7].

Concurrently, the construct of cardiovascular health (CVH) has evolved from Life’s Simple 7 to Life’s Essential 8 and, most recently, to Life’s Crucial 9 (LC9), which adds psychological well-being as a core domain [8–10]. While ideal CVH attenuates CVD risk across CKM stages [11], three critical gaps persist. First, the clinical relevance of the updated LC9 metric within CKM staging is unexplored. Second, whether CVH protection extends uniformly to CVD subtypes with distinct pathobiology, such as heart failure and peripheral artery disease, is unknown. Third, and most fundamentally, the biological mechanisms through which optimal CVH modifies risk at each CKM stage are completely uncharacterized. Without this mechanistic layer, CKM remains a descriptive staging tool rather than a targetable framework.

High-throughput proteomics and metabolomics now enable systematic deconstruction of the molecular architecture linking multisystem disease to clinical endpoints [12–15]. Here, we integrate these platforms with mediation and the LC9 metric in 355,724 UK Biobank participants to: (1) define the stage-specific proteomic and metabolomic signatures of CKM-related CVD risk; (2) map the shifting biological pathways associated with advancing CKM stages; (3) quantify the proportion of the CKM–CVD risk association that is statistically accounted for by these molecules; and (4) test whether the protective association of LC9-defined CVH is statistically accounted for by these same stage-specific molecules. These findings identify stage-specific molecules that statistically account for both CKM-associated risk and CVH-associated protection, providing a molecular framework for future stage-adapted intervention trials.

## Methods

### Study Design and population

We included 355,724 UK Biobank participants free of prevalent CVD at baseline (CKM Stage 4 excluded) **(Analytic Sample 1)**. CKM stages (0-3) were defined per AHA criteria adapted for UK Biobank **(Supplementary Methods)**. Two nested subsamples had proteomic (Olink Explore 3072; n = 37,785) **(Sample 2.1)** [13] and metabolomic (Nightingale NMR; n = 190,112) **(Sample 2.2)** [15] profiling. A subsample of 286,789 participants had complete LC9 data **(Sample 3) (Figure S1)**. **Cardiovascular Health (LC9)**

CVH was quantified using the LC9 score, comprising five health behaviors (diet, physical activity, nicotine exposure, sleep, psychological well-being) and four health factors (blood lipids, glucose, BMI, blood pressure) [8–10]. Each component was scored 0–100 per AHA guidance, with higher scores indicating better health. Dietary scoring followed an established alternate index [16–18]; psychological health was assessed across four domains (depressive symptoms, anxiety, stress, social isolation) [19] **(Supplementary Methods)**. The overall LC9 score was calculated as the unweighted average of all nine components (range 0-100) and categorized as low (<50), moderate (50-79), or high (≥80).

### Outcomes

The primary outcome was incident CVD, a composite of myocardial infarction (MI), stroke, coronary heart disease (CHD), heart failure (HF), atrial fibrillation (AF), and peripheral artery disease (PAD). We also examined each component of this composite as a secondary outcome. Events were ascertained via hospital episode statistics and death registries using ICD-10 codes **(Table S1)** [20,21]. Follow-up extended from baseline to first event, death, loss to follow-up, or censoring.

### Statistical Analysis

#### CKM–CVD associations

Cox proportional hazards models were fitted with sequential adjustment: Model 1 adjusted for age, sex, and ethnicity; Model 2 further adjusted for education, Townsend deprivation index (TDI), employment, household income, and alcohol consumption. Proportional hazards assumptions were verified. Missingness was <1% for all covariates except household income (10.3%). Categorical covariates with <1% missing were imputed by mode, and continuous covariates by mean; household income missingness was modeled as a separate “unknown” category.

#### Multi-omic biomarker selection

For proteomic and metabolomic datasets, Least Absolute Shrinkage and Selection Operator (LASSO) regression identified biomarkers differentiating each progressive CKM stage from Stage 0 (10-fold cross-validation; λ minimizing deviance). To assess feature importance and capture non-linear relationships, we employed Extreme Gradient Boosting (XGBoost) with SHapley Additive exPlanations (SHAP) values on held-out validation sets (70/30 split). Features were ranked by mean absolute SHAP value. XGBoost-SHAP feature importance ranks are descriptive and intended for exploratory feature prioritization; they do not provide formal statistical inference or *P* values for group comparisons.

#### Pathway enrichment

For proteomic data, Gene Ontology (GO) and Kyoto Encyclopedia of Genes and Genomes (KEGG) pathway enrichment analyses were performed. For metabolomic data, enrichment analysis was conducted using MetaboAnalyst 6.0. All enrichment *P*-values were adjusted for multiple comparisons using the Benjamini-Hochberg false discovery rate (FDR) method.

#### Mediation analysis

We performed statistical mediation analysis using the *mediation* R package to quantify the proportion of the exposure-outcome association that was statistically accounted for by each candidate molecule for: (a) CKM stage and incident CVD, and (b) LC9 score and incident CVD, adjusted for Model 2 covariates. For the LC9–CVD mediation analyses, all models were additionally adjusted for CKM stage to account for the shared variance between LC9 and CKM staging criteria. For the outcome model in mediation analyses, we used a Weibull parametric survival model (via *survreg* in R), which is the standard approach within the mediation package framework for survival outcomes. Confidence intervals for mediation effects were estimated using the quasi-Bayesian approximation (1,000 simulations). Given the hypothesis-generating nature of our mediation analyses, we applied FDR correction to control for multiple comparisons across the large number of mediators tested, with statistical significance defined as q<0.05.

#### CVH–CKM interaction

Multiplicative interaction terms in Cox models tested effect modification by CKM stage; restricted cubic splines modeled continuous LC9-CVD associations.

#### Sensitivity analyses

a. further adjustment for CVD polygenic risk score; (b) exclusion of events occurring within the first two years of follow-up; and (c) complete-case analysis excluding participants with any missing covariate data; and (d) performed sensitivity analyses for unmeasured confounding using the E-value approach for mediation analyses [22]; and (e) repeated the joint CKM–CVH analysis using an alternative LC9 score in which psychological health was defined by PHQ-4 instead of ICD-10 codes [9,23].

Detailed definitions and procedures are provided in **Supplementary Methods.** All analyses used R 4.1.1. Two-sided *P* < 0.05 was considered significant; omics analyses applied FDR correction.

## Result

### CKM Staging Defines a Clinical Risk Gradient

Among 355,724 participants (mean age 56.1±8.1 years; 55.5% female), CKM stage distribution was: Stage 0, 13.7%; Stage 1, 12.3%; Stage 2, 73.0%; Stage 3, 1.0% **(Table 1)**. The demographic and clinical characteristics of the proteomic (n = 37,785) and metabolomic (n = 190,112) subsamples were highly comparable to those of the primary analytic sample **(Table S2)**.

**Table 1.** Baseline Characteristics by Cardiovascular-Kidney-Metabolic (CKM) Syndrome Staging Categories *.

| Characteristics | CKM Syndrome Staging |  |  |  | *P* value |
| --- | --- | --- | --- | --- | --- |
|  | Stage 0 | Stage 1 | Stage 2 | Stage 3 |  |
| **N, %** | 48772 (13.7) | 43749 (12.3) | 259614 (73.0) | 3589 (1.0) |  |
| **Age, years** | 52.4 (7.8) | 53.2 (8.0) | 57.2 (7.8) | 64.4 (5.2) | <0.001 |
| **Male, n (%)** | 12907 (26.5) | 15062 (34.4) | 127681 (49.2) | 2757 (76.8) | <0.001 |
| **White, n (%)** | 47112 (96.6) | 41030 (93.8) | 247471 (95.3) | 3285 (91.5) | <0.001 |
| **College or University degree, n (%)** | 22275 (45.7) | 15819 (36.2) | 79942 (30.8) | 715 (19.9) | <0.001 |
| **Employment, n (%)** | 45079 (93.2) | 40714 (93.9) | 240663 (93.5) | 3318 (93.3) | <0.001 |
| **Townsend deprivation index** | -1.5 (2.9) | -1.4 (3.0) | -1.4 (3.0) | -0.6 (3.4) | <0.001 |
| **Household income from 18,000 to 51,999, n (%)** | 21135 (49.0) | 19775 (51.4) | 117452 (52.8) | 1396 (47.5) | <0.001 |
| **Alcohol consumption status, n (%)** |  |  |  |  | <0.001 |
| Never or occasional | 7346 (15.1) | 7739 (17.7) | 49343 (19.0) | 1069 (29.8) |  |
| <1 time/week | 5364 (11.0) | 5399 (12.3) | 28593 (11.0) | 412 (11.5) |  |
| 1–4 times/week | 25986 (53.3) | 22853 (52.3) | 126874 (48.9) | 1402 (39.1) |  |
| Daily or almost daily | 10056 (20.6) | 7732 (17.7) | 54631 (21.1) | 700 (19.5) |  |
| **Smoking status, n (%)** |  |  |  |  | <0.001 |
| Never | 30504 (62.5) | 25739 (58.8) | 141457 (54.5) | 1190 (33.2) |  |
| Previous | 13788 (28.3) | 13836 (31.6) | 91186 (35.1) | 1395 (38.9) |  |
| Current | 4480 (9.2) | 4174 (9.5) | 26971 (10.4) | 1004 (28.0) |  |
| **Body mass index, kg/m<sup>2</sup>** | 22.4 (1.7) | 27.6 (3.2) | 28.1 (4.7) | 30.4 (5.4) | <0.001 |
| **Healthy diet score** | 3.2 (1.4) | 3.1 (1.3) | 3.0 (1.4) | 3.0 (1.5) | <0.001 |
| **Sleep duration, hours/day** | 7.2 (1.0) | 7.1 (1.0) | 7.2 (1.1) | 7.3 (1.3) | <0.001 |
| **Systolic blood pressure, mmHg** | 121.1 (10.4) | 124.0 (9.4) | 143.0 (17.7) | 158.3 (20.9) | <0.001 |
| **Blood cholesterol, mmol/L** | 5.5 (1.0) | 5.6 (1.0) | 5.9 (1.1) | 5.1 (1.2) | <0.001 |
| **Blood Triglycerides, mmol/L** | 1.0 (0.3) | 1.1 (0.3) | 2.0 (1.1) | 2.4 (1.4) | <0.001 |
| **HbA1c, %** | 5.2 (0.3) | 5.3 (0.3) | 5.5 (0.6) | 6.9 (1.7) | <0.001 |
| **Use of antihypertensive medicine, n (%)** | 31 (0.1) | 25 (0.1) | 59174 (22.8) | 2699 (75.2) | <0.001 |
| **Use of cholesterol-lowering medicine, n (%)** | 1073 (2.2) | 1830 (4.2) | 41131 (15.8) | 1812 (50.5) | <0.001 |
| **Life's Crucial 9-Defined Cardiovascular Health, n (%)** |  |  |  |  | <0.001 |
| Low CVH | 28 (0.1) | 276 (0.8) | 12162 (5.9) | 770 (28.4) |  |
| Moderate CVH | 19686 (48.1) | 28720 (80.0) | 182726 (88.2) | 1919 (70.7) |  |
| High (Ideal) CVH | 21184 (51.8) | 6903 (19.2) | 12390 (6.0) | 25 (0.9) |  |

\*Variables are presented as Mean (SD) or n (%).
**Abbreviations:** CKM, cardiovascular-kidney-metabolic syndrome; CVH, Cardiovascular Health.

During median 13.5-year follow-up, 53,587 incident CVD events occurred. CVD risk increased generally with advancing CKM stage. In fully adjusted models, hazard ratios (HRs) versus Stage 0 were: Stage 1, 1.17 (95% CI, 1.12-1.23); Stage 2, 1.78 (1.72-1.85); Stage 3, 3.51 (3.31-3.73) (*P*-trend < 0.001). Associations were steepest for heart failure (Stage 3, HR, 6.77 [5.94-7.71]) and peripheral artery disease (7.54 [6.29-9.02]) **(Figure S2)**; complete subtype-specific hazard ratios are provided in **Table S3**. Results were robust to further adjustment for polygenic risk score, exclusion of events within the first two years, and complete-case analysis **(Table S4)**.

### Stage-Specific Multi-Omic Signatures of CKM–CVD Relationships

LASSO regression identified 11 pan-stage proteins consistently dysregulated across Stages 1-3, including reticulon-4 receptor (RTN4R), leptin (LEP), low-density lipoprotein receptor (LDLR), and adipocyte fatty acid-binding protein (FABP4), and 29 pan-stage metabolites, including glycoprotein acetyls (GlycA), linoleic acid, glutamine, and creatinine **(Figure 1a, 1b)**. Venn analysis revealed both shared and unique molecular architectures **(Figure 1a, 1b)**, with stage-specific and transitional signatures visualized in **Figure 1e, 1f** (proteins) and **Figure 1i, 1j** (metabolites). Full lists of LASSO-selected proteins and metabolites for each stage comparison are provided in **Tables S5–S10**. The results of category distribution, functional enrichment, and PPI networks for each stage are presented in **Figures S3–S4**; descriptive network metrics are provided in **Table S11**.

**Figure 1.**
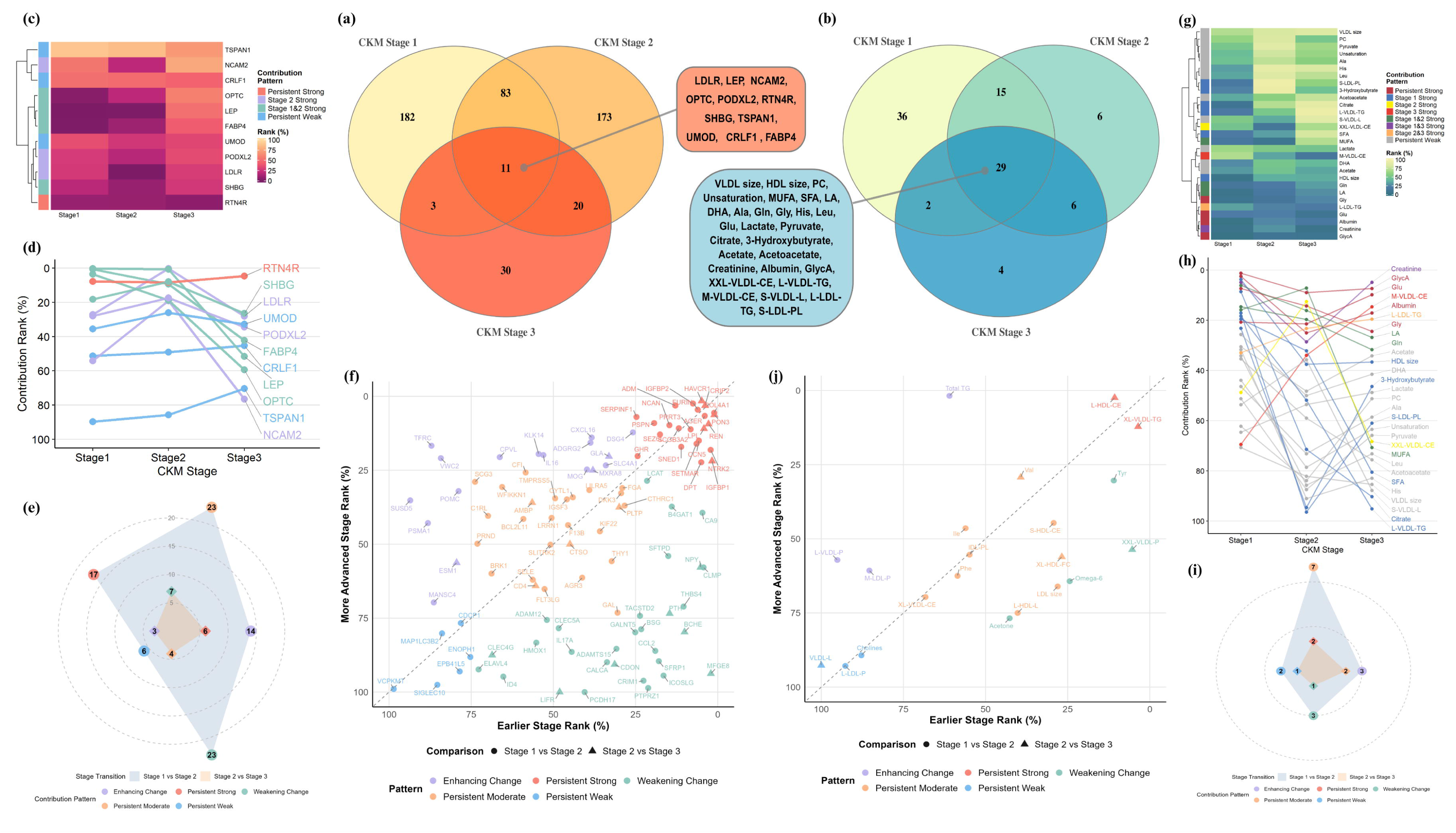
Stage-Resolved Dynamics of Proteomic and Metabolomic Signatures Across CKM Stages. ***CKM stages are characterized by both persistent and dynamically shifting molecular signatures, with stage-specific biomarker relevance revealed by XGBoost-SHAP.*** **Panels a and b (LASSO-selected biomarkers).** Venn diagrams show overlapping and unique proteins (a) and metabolites (b) associated with each progressive CKM stage (Stage 1–3 *vs.* Stage 0). **Panels c and d (Contribution dynamics of pan-stage proteins).** Heatmap (c) depicts SHAP-based relative importance; line plot (d) tracks ranking trajectories. RTN4R shows sustained strong importance; FABP4 and LEP dominate early stages but decline in Stage 3; LDLR exhibits Stage 2-specific prominence. **Panels e and f (Transition patterns of stage-shared proteins).** Radar plot (e) shows the count distribution of five contribution patterns—Enhancing Change, Weakening Change, Persistent Strong, Persistent Moderate, and Persistent Weak—for the Stage 1–to–2 (blue) and Stage 2–to–3 (orange) transitions; radial distance indicates count. Scatter plot (f) plots each protein’s rank percentile in the earlier stage (x-axis) against its rank in the later stage (y-axis); points above the diagonal indicate improved rank with stage progression. **Panels g and h (Contribution dynamics of pan-stage metabolites).** Heatmap (g) and ranking trajectories (h) show persistently strong GlycA, Stage 1-dominant HDL particle size, Stage 2-peaked cholesteryl esters in chylomicrons/VLDL, and dual-stage dominance of creatinine (Stages 1 and 3). **Panels i and j (Transition patterns of stage-shared metabolites).** Radar plot (i) and scatter plot (j) follow the same format as panels e and f, respectively, illustrating parallel trajectory patterns for metabolites. **Abbreviations:** CVD, cardiovascular disease; LASSO, least absolute shrinkage and selection operator; SHAP, Shapley additive explanations; XGBoost, extreme gradient boosting; HR, hazard ratio.

### Stage-Specific Contribution Patterns of Biomarkers

XGBoost-SHAP revealed that the relevance of specific biomarkers changes dynamically with stage **(Figure 1c, 1d [proteins], 1g, 1h [metabolites])**. Among pan-stage proteins, RTN4R maintained consistently strong importance across all stages. In contrast, FABP4 and LEP dominated early stages but declined markedly in Stage 3, while LDLR exhibited Stage 2-specific prominence.

Among pan-stage metabolites, GlycA showed persistently strong contributions. HDL particle size dominated Stage 1; cholesteryl esters in chylomicrons/very large VLDL peaked in Stage 2; creatinine showed dual-stage dominance (Stages 1 and 3). Total triglycerides emerged as the top contributor in Stage 2, marking a metabolic shift. Complete SHAP feature importance rankings are provided in **Tables S12-S13** and **Figures S5–S6**.

Radar plots **(Figure 1e, 1i)** and scatter plots **(Figure 1f, 1j)** illustrate how the relative contribution of stage-shared molecules reconfigures across transitions. The Stage 1–to–2 transition was dominated by weakening change and persistent moderate patterns (23 each), whereas the Stage 2–to–3 transition showed substantially lower counts across all categories, indicating a narrowing molecular landscape as CKM advances. Scatter plots **(Figure 1f, 1j)** further show that early adiposity-related signals (e.g., FABP4, LEP) declined in rank, while later fibrotic/renal signals (e.g., Hepatitis A virus cellular receptor 1 [HAVCR1], creatinine) gained prominence.

### Pathway Dynamics: From ECM to Inflammation to Fibrosis

Pathway enrichment delineated a coherent, stage-ordered trajectory of biological dysregulation **(Figure 2a, 2b)**. This trajectory supports the biological coherence of the CKM staging construct at the pathway level. Stage 1 was characterized by extracellular matrix (ECM) reorganization, vesicle transport, and branched-chain amino acid biosynthesis. Stage 2 featured inflammatory amplification (cytokine– cytokine interactions, MAPK/ERK signaling), steroidogenesis, and bile acid metabolism. Stage 3 was defined by hypoxia response, antibacterial defense, and fibrotic remodeling.

**Figure 2.**
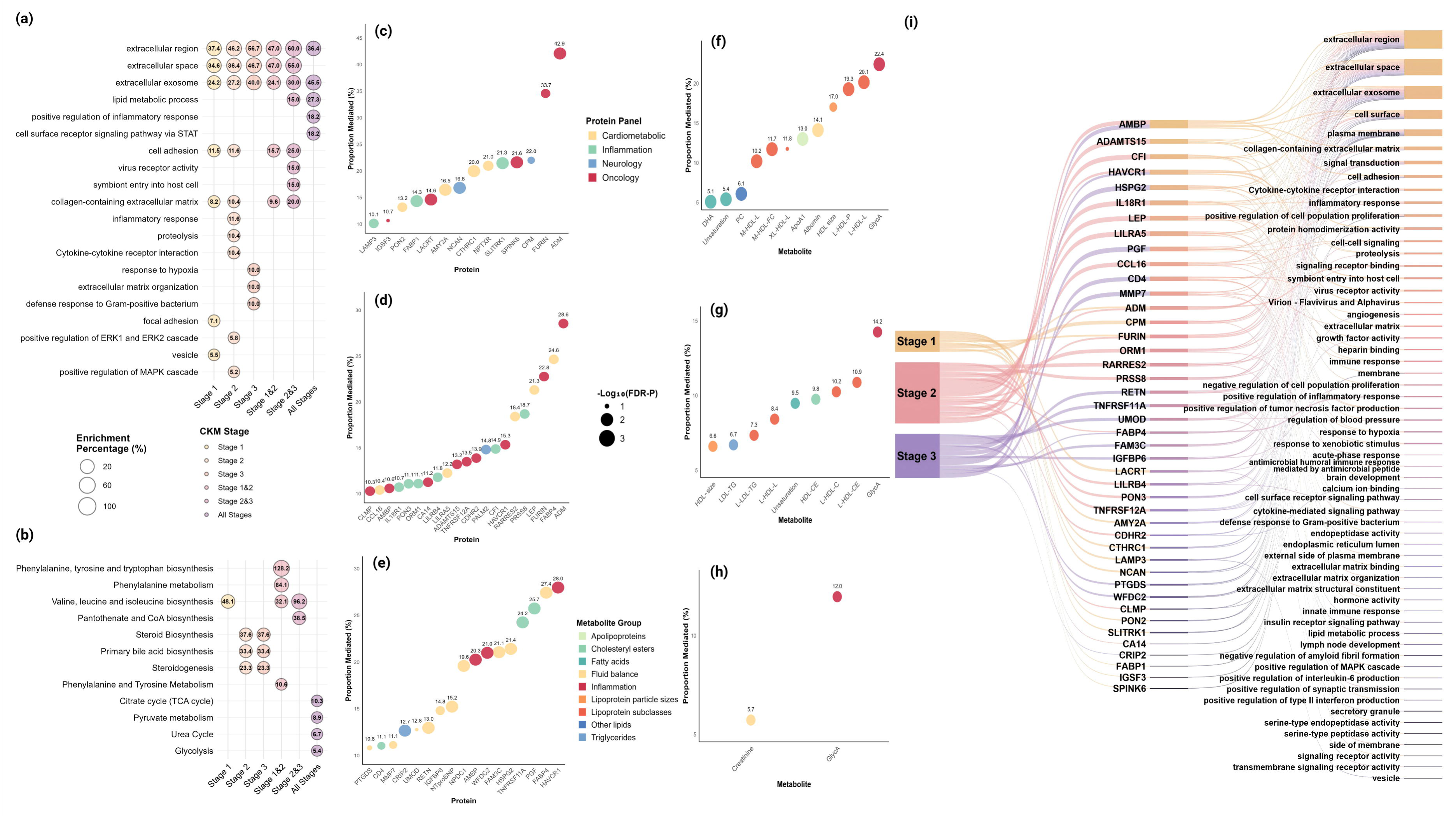
Multi-Omic Landscape of CKM Stages and CVD Risk Mediation. ***CKM stages are associated with a stage-specific pattern of biological dysregulation, with relative enrichment of ECM/metabolic pathways in Stage 1, inflammatory pathways in Stage 2, and fibrotic/hypoxic pathways in Stage 3. Stage-specific multi-omic mediators quantify the contribution of these pathways to excess CVD risk.*** **Panels a and b (Pathway dynamics across CKM stages).** Panel a shows proteomic pathway enrichment; **Panel b** shows metabolomic pathway enrichment. Circle size represents the gene ratio (a) or enrichment ratio (b); color intensity indicates statistical significance (FDR-adjusted P-value). Full pathway lists are provided in Tables S14– S15. **Panels c through i (Stage-specific mediation of CKM–CVD risk). Panels c, d, and e** present proteomic mediators for CKM stages 1, 2, and 3, respectively. **Panels f, g, and h** present metabolomic mediators for CKM stages 1, 2, and 3, respectively. **Panel i** provides a cross-comparison of stage-specific mediators. Forest plots display proportion mediated with 95% confidence intervals. Mediation analyses were adjusted for multiple comparisons using FDR correction (q<0.05). All mediation models were adjusted for age, sex, ethnicity, education, Townsend deprivation index, employment, household income, and alcohol consumption. All mediation analyses are cross-sectional and do not establish causality. Full results are provided in Tables S16– S17.

Analysis of transition interfaces revealed coordinated molecular adaptations between stages. The Stage 1–to–2 transition was marked by enhanced cell adhesion mechanisms (proteomic) and aromatic amino acid metabolism (metabolomic), suggesting remodeling of cell–matrix interactions and metabolic reprogramming as key drivers of early progression. The Stage 2–to–3 transition featured induction of viral entry pathways (proteomic)—implicating endosomal trafficking dysfunction and innate immune dysregulation—alongside enhanced cofactor synthesis (metabolomic).

Collectively, these findings delineate a stage-associated pattern in which ECM remodeling and metabolic pathways are relatively more enriched in Stage 1, inflammatory pathways in Stage 2, and fibrotic/hypoxic pathways in Stage 3, with substantial overlap across adjacent stages. Full pathway enrichment results, including all GO terms and KEGG pathways with FDR-corrected *P*-values, are provided in **Tables S14–S15**.

### Stage-Specific Mediators of the CKM–CVD Risk Association

Mediation analysis demonstrated a fundamental shift in the molecular drivers of CVD risk across CKM stages **(Figure 2c–2i)**. This shift indicates that the molecules statistically accounting for CKM–CVD risk associations are largely stage-specific, consistent with distinct biological processes operating at each phase of CKM progression. In Stage 1, a substantial proportion of the CKM-CVD association was statistically accounted for by ECM remodeling proteins—pro-adrenomedullin (ADM, 42.9%), furin (FURIN, 33.7%), carboxypeptidase M (CPM, 22.0%)—and lipoproteins (GlycA, 22.4%). In Stage 2, the profile shifted to inflammatory mediators—FABP4 (24.6%), LEP (21.3%), and prostasin (PRSS8, 18.7%). In Stage 3, renal injury and fibrotic markers predominated—HAVCR1 (28.0%), placenta growth factor (PGF, 25.7%), and WAP four-disulfide core domain protein 2 (WFDC2, 21.0%). GlycA functioned as a pan-stage molecule that statistically accounted for a significant portion of the association. Complete mediation results for all tested proteins and metabolites are provided in **Tables S16-S17**. Due to the limited number of events in Stage 3, the confidence intervals for some estimates are wide and should be interpreted cautiously. Sensitivity analyses using the E-value approach **(Table S18)** suggested that the observed indirect effects were robust to potential unmeasured confounding.

To move beyond individual mediators and provide a more comprehensive pathway-level view, we performed enrichment analysis on the full set of main significant mediators identified in each stage (**Tables S19-S20**, proportion of mediation >10%). In Stage 1, mediator-enriched proteins were predominantly localized to the extracellular region and cell surface (e.g., CPM, ADM, FURIN). Metabolomic enrichment highlighted alpha-linolenic and linoleic acid metabolism, biosynthesis of unsaturated fatty acids, and phenylalanine/tyrosine/tryptophan metabolism, consistent with early metabolic dysregulation and lipid remodeling. In Stage 2, the mediator protein landscape shifted toward cell surface and extracellular exosome compartments, with significant enrichment of inflammatory response pathways, cytokine-cytokine receptor interaction, and serine-type peptidase activity; metabolomic enrichment identified amino sugar and nucleotide sugar metabolism. In Stage 3, mediator proteins were enriched in extracellular region and cell surface, with functional themes including defense response, lipid binding, and inflammatory response; metabolomic enrichment consistently implicated amino sugar and nucleotide sugar metabolism. These pathway-level patterns support a stage-associated shift in biological emphasis—from ECM/metabolic disturbances in Stage 1, through inflammatory signaling in Stage 2, to fibrotic/hypoxic responses in Stage 3—while acknowledging that individual mediators within each stage contribute to overlapping pathways.

Detailed CVD subtype-specific mediation results are provided in supplementary results **(Tables S21-S22, Figures S7–S9)**.

### Stage-Dependent Protection of LC9-Defined CVH

CVH deteriorated progressively with advancing CKM stage **(Figure 3a, S10)**. The protective association of high CVH was strongly modified by CKM stage (*P*-interaction = 0.012). Compared to the reference (Stage 3, low CVH), high CVH (LC9≥80) was associated with 81% lower CVD risk in Stage 0 (adjusted HR, 0.19; [95% CI, 0.17-0.22]), 79% lower risk in Stage 1 (0.21 [0.18-0.24]), and 72% lower risk in Stage 2 (0.28 [0.25-0.31]). In Stage 3, the estimate was highly imprecise (0.86, [0.44–1.66]) **(Figure 3b)**, owing to only 25 participants with high CVH; no reliable inference could be drawn for this subgroup. Sensitivity analyses using an alternative LC9 score with PHQ-4-based psychological health definition yielded consistent results **(Table S4)**.

**Figure 3.**
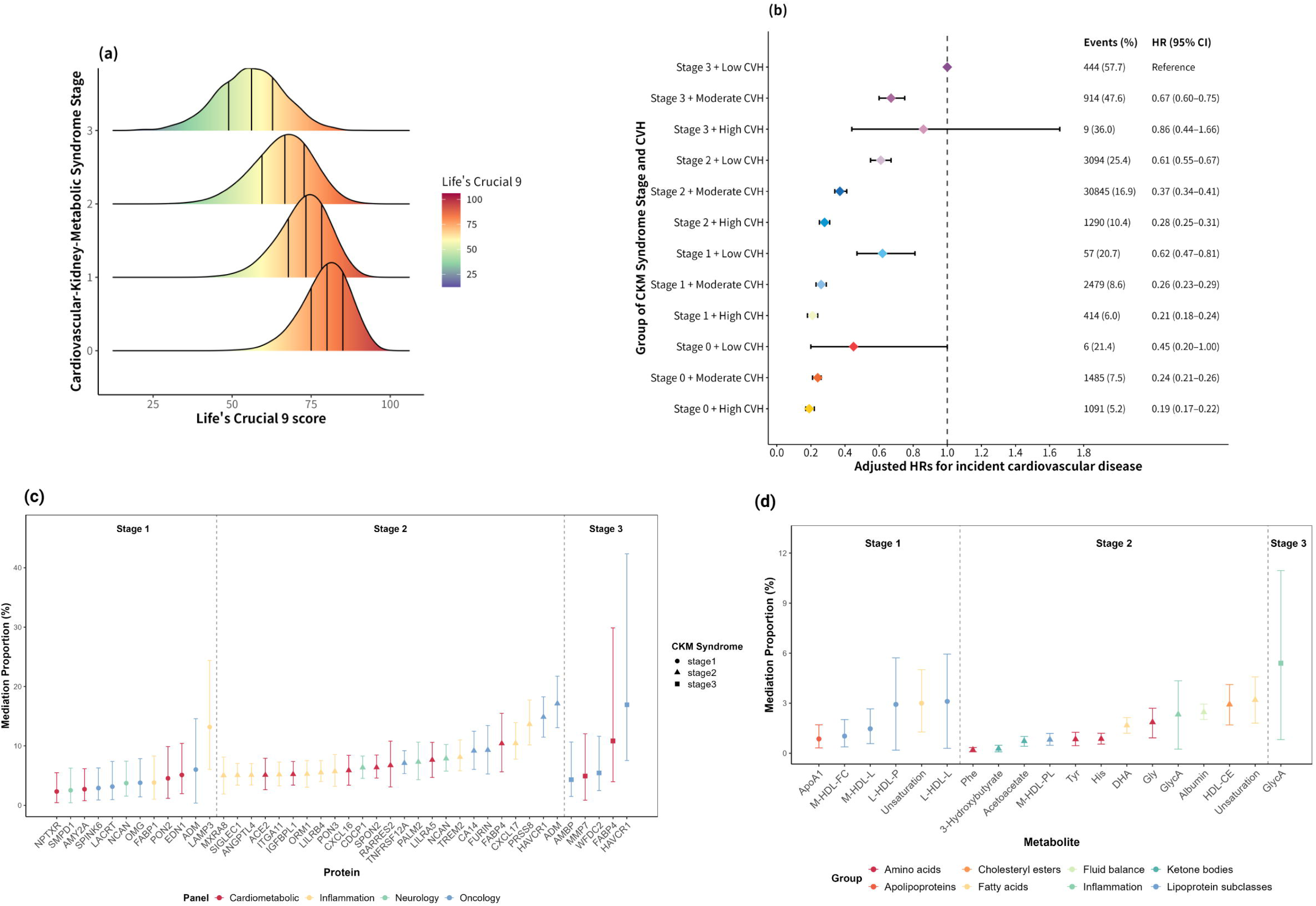
Stage-Dependent Protection of Cardiovascular Health (LC9) Against CVD Risk Across CKM Stages*. ***CVH protection shows strong stage-dependent associations and is statistically accounted for by the same stage-specific molecules associated with CKM pathology.* Panels a and b (Distribution and joint associations). Panel** **a** presents the LC9 score distribution across CKM stages (ridge plot). **Panel b** shows the joint associations of CKM stage and CVH with CVD risk (forest plot). **Panels c and d (Mediation analyses). Panel c** displays protein mediators of the LC9–CVD association (forest plot). **Panel d** displays metabolite mediators of the LC9–CVD association (forest plot). *Forest plots display hazard ratios (Panel b) or proportion mediated (Panels c and d) with 95% confidence intervals. Analyses were adjusted for age, sex, ethnicity, education, Townsend deprivation index, employment, household income, and alcohol consumption, and (for LC9–CVD mediation analyses only) additionally for CKM stage. All mediation analyses are cross-sectional and do not establish causality. Only features significantly mediating both CKM–CVD and LC9–CVD pathways are shown. Full results in Tables S23–S24.

### Multi-Omic Mediators of CVH Protection

In mediation analysis, a proportion of the protective association of LC9 with CVD was statistically accounted for by the same stage-specific molecules that accounted for the CKM–CVD risk association **(Figure 3c, 3d)**. These findings are consistent with the hypothesis that some of the protective mechanisms of ideal CVH may involve attenuation of the same pathways that drive CKM-stage-specific CVD risk. In Stage 1, a proportion of the protective association was statistically accounted for by lysosome-associated membrane glycoprotein 3 (LAMP3, 13.2%), ADM (6.0%), and serum paraoxonase/arylesterase 2 (PON2, 4.5%). In Stage 2, ADM (17.1%), HAVCR1 (14.9%), and PRSS8 (13.7%) were the predominant mediators. In Stage 3, HAVCR1 (16.9%) and GlycA (5.4%) accounted for the residual protective signal. Cross-stage analysis identified five consistent mediators of CVH protection: ADM, FABP4, HAVCR1, fatty acid unsaturation degree, and GlycA. Complete LC9 mediation results are provided in **Tables S23-S24**.

## Discussion

This large-scale study provides the first stage-resolved molecular atlas of CKM-related CVD risk. By integrating deep multi-omics with mediation analysis and the LC9 metric in 355,724 UK Biobank participants, we make three observations. First, CKM stages are associated with a stage-ordered molecular continuum—from ECM remodeling and metabolic dysregulation (Stage 1) to inflammatory amplification (Stage 2) and fibrotic/renal injury (Stage 3)—that delineates the shifting stage-associated drivers of CVD risk. Second, the protective efficacy of ideal CVH (LC9 ≥80) is strongly stage-dependent: ∼80% risk reduction in early Stages, whereas in Stage 3 the estimate was highly imprecise owing to only 25 participants with high CVH, precluding definitive interpretation. Third, in mediation analysis, a proportion of this protective association was statistically accounted for by the same stage-specific molecules that statistically account for CKM–CVD risk association. This convergence—where the same molecules statistically account for both CKM-associated CVD risk and LC9-associated protection—generates the hypothesis that these molecules lie on a modifiable pathway. These findings are hypothesis-generating and provide a rationale for future intervention studies.

### The CKM Staging Framework: A Molecular and Clinical Gradient

Our findings molecularly substantiate the CKM clinical risk gradient [4,24]. The graded increase in CVD risk is underpinned by an evolving molecular continuum—from adipocentric disturbances (LEP, FABP4) in early stages to hypoxia, fibrotic, and renal injury signatures (HAVCR1, PGF) in Stage 3 [25,26]. The strong role of CKM Stage 3 reflects the cumulative burden of risk factors captured by the CKM staging criteria, rather than an effect independent of its components. Mediation analysis further quantifies the contribution of these stage-specific molecules: they account for 18–43% of the excess CVD risk conferred by each CKM stage. This suggests that the CKM staging construct is associated with distinct biological processes that evolve with disease stage, supporting its construct validity.

Notably, the CKM–CVD risk gradient was most pronounced for heart failure and peripheral artery disease (Stage 3 HRs: 6.77 and 7.54, respectively), suggesting that CKM pathophysiology exerts a particularly potent impact on vascular and myocardial end-organ vulnerability. The subtype-specific patterns observed in supplementary analyses warrant mechanistic exploration to understand how different CVD subtypes diverge in their responsiveness to CKM-related pathophysiology.

### Stage-Resolved Pathways and Preclinical Evidence for the CKM Continuum

Our pathway analysis maps a stage-associated pattern of biological dysregulation, with relative enrichment of ECM remodeling among Stage 1, inflammatory signaling among Stage 2, and fibrotic/hypoxic responses among Stage 3 [27,28]. This map identifies potential biologically salient pathways within the CKM continuum. The prominence of cytokine-related signaling (IL-6/JAK/STAT) and GlycA in Stage 2, for example, identifies inflammation as the dominant potential driver at this transition [29]. Our mediation analysis further quantifies the extent to which these molecules statistically account for both the CKM–CVD and LC9–CVD associations. This convergent evidence moves pathway analysis from descriptive observation toward hypothesis generation for targeted intervention.

The convergence between our findings and recent preclinical evidence further strengthens this mechanistic framework. Bollinger et al. recently demonstrated that restoration of branched-chain amino acid (BCAA) catabolism improves kidney function in preclinical CKM models, implicating BCAA dysregulation in early CKM pathogenesis [30]. Our observation that BCAA biosynthesis is selectively enriched in Stage 1 directly extends this experimental evidence to the human population level, providing complementary support for BCAA metabolism as an early potential targetable node in the CKM continuum.

However, our cross-sectional design precludes quantification of dynamic pathway interactions. Future longitudinal studies employing network physiology frameworks [31]—using centrality metrics or perturbation analyses on serial multi-omics data—could address this and identify transition points not apparent from static enrichment alone.

### LC9 is Associated with Stage-Specific Molecular Architecture

The central finding of this study is that the protective association of LC9-defined CVH is statistically accounted for by the same stage-specific molecules that account for CKM-associated CVD risk. This pattern is consistent with, but does not prove, a causal pathway. Because LC9 was measured observationally at baseline and not experimentally manipulated, we cannot establish causality.

In **Stage 1**, the protective association is statistically accounted for by ADM and LAMP. ADM is a well-established mediator in vascular pathways [32], while LAMP3 has been implicated in extracellular matrix remodeling processes relevant to tissue integrity [33]. LC9 health factors (blood pressure, lipids, glucose) directly target these pathways, while health behaviors (diet, physical activity, sleep) likely influence them indirectly via weight regulation and metabolic control.

In **Stage 2**, the protective association is statistically accounted for through inflammatory/renal molecules (HAVCR1, PRSS8) [34]. Here, physical activity, sleep, and psychological health become the dominant levers, alongside lipid and glucose control.

In **Stage 3**, the residual protective signal is statistically accounted for by HAVCR1 and GlycA, consistent with the persistence of renal injury and inflammatory pathways in advanced disease [35,36]. However, the estimate of the LC9–CVD association in Stage 3 was based on only 25 participants with high CVH, yielding a wide confidence interval (0.44–1.66) that cannot distinguish between no effect and meaningful protection. This precludes conclusions about the role of CVH in Stage 3 and underscores the need for larger studies.

These observations suggest that LC9 may function not merely as a risk marker, but as a health metric associated with stage-specific molecular pathology; however, this observation is associative and requires experimental validation. CKM Stage 1 and Stage 2 criteria intentionally overlap with LC9 health factors (BMI, blood pressure, glucose), reflecting the convergent architecture of cardiometabolic risk. All LC9– CVD mediation models were adjusted for CKM stage, ensuring that the identified mediators reflect pathways beyond these shared definitions.

### Clinical and Translational Synthesis: From Mechanism to Actionable Strategy

The convergence of our molecular map and LC9 findings generate hypotheses for a stage-adapted precision prevention paradigm.

First, dynamic biomarker-guided risk assessment. Pan-stage (GlycA) and stage-specific (HAVCR1) molecules may serve as candidate biosensors of pathological activity, with potential utility for monitoring intervention efficacy and refining risk assessment beyond traditional factors—pending prospective validation.

Second, stage-adapted intervention hypotheses. Our data generate the hypothesis that intervention priorities may need to shift across stages: intensive metabolic/vascular management in Stages 0–1 to counter early ECM dysregulation; reinforced focus on physical activity, sleep, and psychological health in Stage 2 to address inflammatory drivers; and emphasis on renal protection in Stage 3. Because the role of CVH in Stage 3 could not be reliably assessed in this study, pharmacologic strategies (SGLT2i, GLP-1 RA) remain essential in advanced stages as recommended by current guidelines.

Third, LC9 as an observational platform for hypothesis generation. The LC9 score integrates health behaviors and factors that may influence the stage-specific pathways we identified. While our findings are observational, they suggest testable hypotheses for future lifestyle intervention studies.

### Limitations

Several limitations should be considered. First, our stage-associated molecular signatures are cross-sectional, as proteins, metabolites, and CKM stage were all measured at baseline; we cannot determine whether these molecules predict or reflect within-individual progression. Serial CKM staging and repeated multi-omics are needed for validation. Second, residual confounding is possible; however, E-value sensitivity analyses supported the robustness of our mediation results to unmeasured confounding. Third, the UK Biobank is predominantly White; validation in diverse cohorts is required. Fourth, because exposure and mediators were measured concurrently, our mediation analyses cannot establish temporal precedence or causality and should be interpreted as hypothesis-generating. Fifth, the Stage 3 high CVH analysis was based on only 25 participants, limiting precision and precluding definitive interpretation. Sixth, LC9 was measured observationally, not experimentally manipulated; randomized controlled trials are needed to test the causal hypotheses generated by this study.

### Conclusion

In conclusion, this hypothesis-generating study identifies stage-specific proteomic and metabolomic signatures that statistically account for both CKM-associated CVD risk and the protective association of CVH. These findings generate the hypothesis that CVH may exert its benefits in part through modulation of these molecules. Future randomized trials are needed to test whether modifying these mediators causally reduces CVD risk and whether improving CVH specifically targets these pathways.

## Data Availability

All data produced in the present study are available upon reasonable request to the authors.

## Declarations

### Ethics approval and consent to participate

The UK Biobank study received ethical approval from the NHS National Research Ethics Service (11/NW/0382), and all participants provided written informed consent in accordance with the Declaration of Helsinki.

### Consent for publication

Not applicable.

### Availability of data and materials

The data are available on application to the UK Biobank (https://www.ukbiobank.ac.uk), and the analytic methods and study materials that support the findings of this study will be available from the corresponding authors on request.

### Statement on AI Use

During the preparation of this manuscript, the authors used generative AI tools for language polishing and grammar refinement. All content was reviewed, edited, and approved by the authors, who take full responsibility for the final manuscript.

## Acknowledgments

The authors would be willing to thanks the UK Biobank participants.

